# Machine learning for ovarian cancer: lasso regression-based predictive model of early mortality in patients with stage I and stage II ovarian cancer

**DOI:** 10.1101/2020.05.01.20088294

**Authors:** Robert Chen

**Affiliations:** Emory University School of Medicine

**Keywords:** ovarian cancer, testis, genital cancer, machine learning

## Abstract

While machine learning has shown promise in prediction of mortality in situations such as intensive care units, there is limited evidence of its application towards ovarian cancer.

In this study, we extracted clinical covariates from a cohort of 273 patients with stage I and II ovarian cancer, and trained a machine learning algorithm, L2-regularized logistic regression, on the set of patients in prediction problem for mortality less than 20 months, representing the 25th percentile of overall survival.

Our model achieves an AUC of 0.621, accuracy 0.761, sensitivity 0.130, positive predictive value 0.659, and F1 score 0.216. This study serves as a proof of concept for a predictive model customized towards mortality prediction for malignant neoplasm of the left testis, and can be adapted and generalized to related tumors such as spermatic cord and scrotal tumor types.

## 1. INTRODUCTION

In 2020, there are estimated to be 21,750 new cases of ovarian cancer, representing 1.2% of all new cancer cases. Furthermore, there are estimated to be 13,940 deaths in 2020 from ovarian cancer, representing 2.4% of all cancer-related deaths. While ovarian cancer has an overall 5-year relative survival rate of 48.6%, it is imperative that proper management be conducted at early stages of the disease course.

While there does not exist an intuitive method for risk stratification of patients with ovarian cancer, there is strong evidence of the potential of machine learning for stratification of patients in other diseases such as heart failure (Rasmy et al. 2018; Choi et al. 2017; Chen et al. 2019; Ng et al. 2017), kidney disease (Makino et al. 2019), and critical care(Chen, Su, et al. 2015; Chen, Kumar, et al. 2015; Katuwal and Chen 2016; Kamat et al. 2018; Yu, Liu, and Nemati 2019; Johnson et al. 2016). Furthermore, machine learning has been shown to be effective for readmission prediction (Rajkomar et al. 2018; Desautels et al. 2017; Chen, Su, et al. 2015), drug adverse event prediction (Cheng and Zhao 2014). While such a method does not exist, we posit that a machine learning-based method can be useful for clinical decision support in the management of ovarian cancer. Despite the fact that there is no established risk stratification protocol for ovarian cancer, there exists intuitive sense that stratification may be made possible via machine learning based methods.

In this study we developed and fine-tuned a machine learning model based on L2 regularized logistic regression for prediction of early mortality in patients with ovarian cancer from retrospective real-world data and evaluated their performance. Our model leverages predictive features in a variety of domains including demographics, histology, staging, tumor spread and metastatic status.

## 2. METHODS

A cohort of patients was selected from the The Surveillance, Epidemiology and End Results (SEER) program public retrospective dataset(Hankey, Ries, and Edwards 1999).

### 2.1 Cohort Construction

The Surveillance, Epidemiology and End Results (SEER) program was used to identify a cohort of 85,582 female patients who were diagnosed with ovarian cancer meeting the ICD9 code criteria: C569 - Malignant neoplasm of unspecified ovary. Of these patients, we filtered down a cohort of 14,962 patients whose initial diagnosis date was between 2010 and 2016. Of these patients, 4,202 patients met staging criteria of stage I or II. Finally, we filtered out patients lost to follow-up resulting in a final cohort of 4,201 patients. Table 1 shows descriptive statistics of the cohort.

**Table 1:** Baseline characteristics of the study cohort.^1^,^2^

|  | Survival < 2 mos | Survival >= 22 mos | Total |
| --- | --- | --- | --- |
| **Characteristic** | *n* = 1078 | *n* = 3140 | *N* = 4128 |
| **Age (mean)** | 58.9 | 54.3 | 55.5 |
| **Sex (% female)** | 100% | 100% | 100% |
| **Tumor Size** | 2.73cm | 2.99cm | 2.92cm |
| **Race (%)** |  |  |  |
| American Indian/Asian | 13.3% | 11.9% | 12.3% |
| Black | 7.5% | 7.3% | 7.3% |
| White | 78.4% | 80.0% | 79.6% |
| Hispanic | 10.4% | 8.6% | 9.0% |
| **Group Stage (%)** |  |  |  |
| I | 4.4% | 3.3% | 3.6% |
| IA | 35.3% | 42.1% | 40.4% |
| IB | 39.0% | 5.2% | 48.6% |
| IC | 24.4% | 28.1% | 27.1% |
| II | 0.19% | 0.22% | 0.21% |
| II NOS | 0.11% | 0.22% | 0.59% |
| IIA | 6.4% | 5.6% | 5.8% |
| IIB | 14.0% | 8.1% | 9.6% |
| IIC | 10.4% | 7.0% | 7.9% |
| **Histology (%)** |  |  |  |
| unspecified neoplasms | 2.4% | 0.4% | 0.9% |
| epithelial neoplasms, NOS | 4.5% | 2.2% | 2.8% |
| complex epithelial neoplasms | 0.6% | 0.4% | 0.5% |
| specialized gonadal neoplasms | 3.2% | 5.3% | 4.7% |
| nevi and melanomas | 0.2% | 0.4% | 0.3% |
| squamous cell neoplasms | 3.8% | 3.3% | 3.4% |
| complex mixed and stromal neoplasms | 4.1% | 1.6% | 2.2% |
| fibroepithelial neoplasms | 0.3% | 0.2% | 0.2% |
| germ cell neoplasms | 3.7% | 6.0% | 5.4% |
| trophoblastic neoplasms | 0.0% | 0.0% | 0.0% |
| mesonephromas | 0.0% | 0.0% | 0.0% |
| basal cell neoplasms | 0.6% | 1.1% | 1.0% |
| transitional cell papillomas and carcinomas | 0.2% | 0.4% | 0.3% |
| adenomas and adenocarcinomas | 43.0% | 45.1% | 44.6% |
| adnexal and skin appendage neoplasms | 0.0% | 0.1% | 0.1% |
| cystic, mucinous and serous neoplasms | 33.1% | 33.3% | 33.3% |
| ductal and lobular neoplasms | 0.4% | 0.3% | 0.3% |
| **Metastases (%)** |  |  |  |
| Bone | 0% | 0% | 0% |
| Brain | 0% | 0% | 0% |
| Liver | 0% | 0% | 0% |
| Lung | 0% | 0% | 0% |

Features corresponding to several domains were extracted for the cohort. These include demographics, AJCC staging criteria, and metastatic sites.. Table 2 shows the description of features used in the model.

**Table 2:**
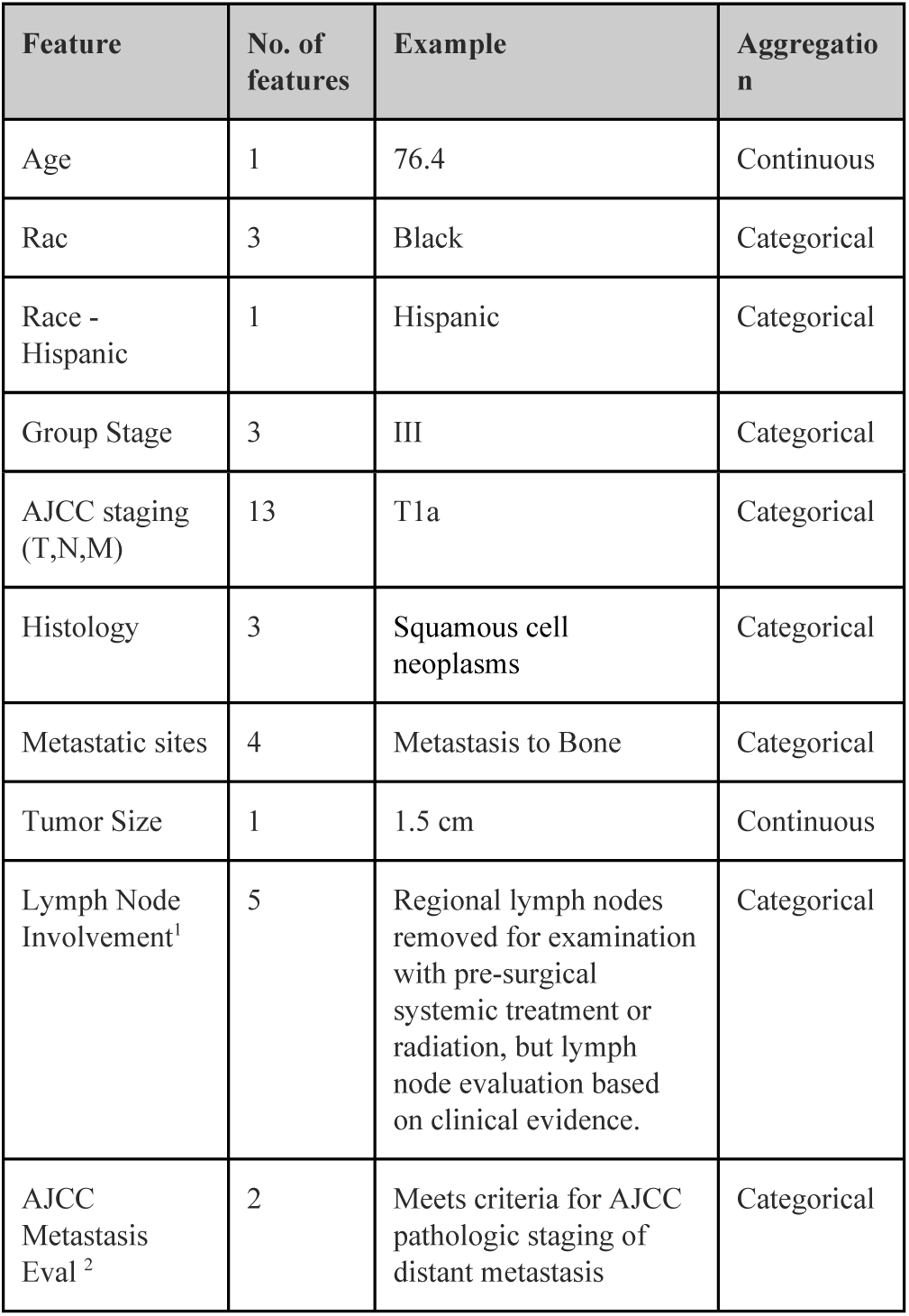
Features used in the model, as well as aggregation used in data preprocessing before model training.

### 2.2 Kaplan Meier Analysis

A Kaplan Meier(Kaplan and Meier 1958) analysis was performed with all patients in the cohort. The 25th percentile of overall survival time in months, was used as a threshold to determine class labels of patients:

- 0: patient survived at least the 25th percentile of overall survival
- 1: patient death occurred before the 25th percentile of overall survival

### 2.2. Machine Learning Model

#### 2.1.1 Feature Construction

Categorical features were one-hot encoded into separate features. For example, the feature race which includes categories (white, black, other) would be one hot encoded for a patient as (1,0,0) for white, (0,1,0) for black, and (0,0,1) for other. Continuous features were used in the current form. Standardization was performed on all features.

#### 2.1.2 Predictive Modeling

Principal component analysis was performed on the cohort to reduce dimensionality. Feature selection was performed using ANOVA F-value for the. A logistic regression (Hastie, Tibshirani, and Friedman 2009) model was trained using the features constructed, with the target labels:

0: patient survived at least the 25th percentile of overall survival

1: patient death occurred before the 25th percentile of overall survival.

Grid search was performed to learn the most optimal set of modeling parameters from the following set: number of features {all}, regularization {l1, l2}, C {1e-2, 1e-1, 1, 1e1, 1e2}. The model was evaluated via 5-fold cross validation. The scikit-learn 24 Python package was used to implement the analysis.

## 3. RESULTS

### 3.1 Kaplan Meier Analysis

The median overall survival was 40 months. The 25th percentile of survival time was 22 months, which was used in the definition of patient classes for the machine learning problem (Figure 1).

**Figure 1:**
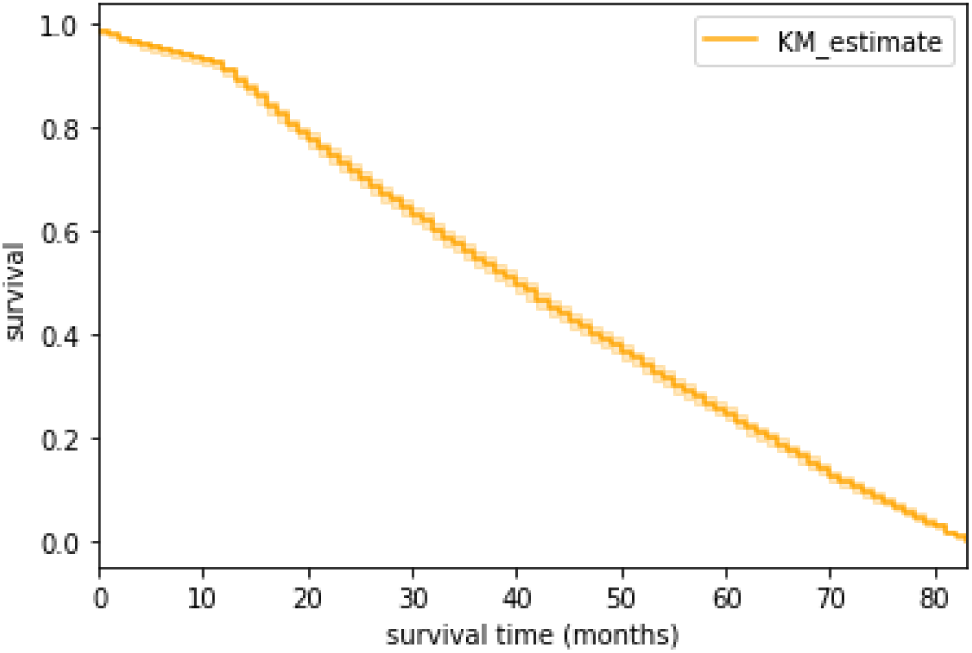
Kaplan Meier survival curve of all patients in the analytical cohort.

### 3.2 Principal Component Analysis

We utilized principal components analysis (PCA) to visualize the variation in the cohort of patients. Figure 2 shows the patients projected onto the first two principal components.

**Figure 2:**
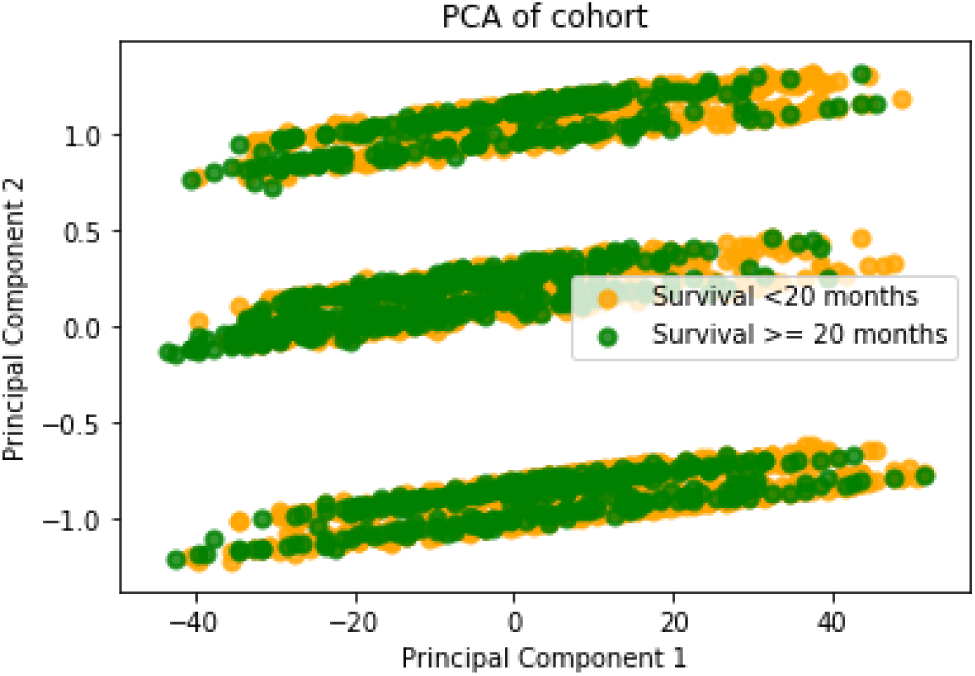
PCA plot of patients. Patients surviving less than 11 months are shown in red; otherwise green.

### 3.3 Predictive Model Performance Metrics

Across all folds of cross validation, the model achieved AUC of 0.621, accuracy of 0.761, precision of 0.659, recall of 0.130, and F1 score 0.216. The receiver operating curve is shown in figure 3.

**Figure 3:**
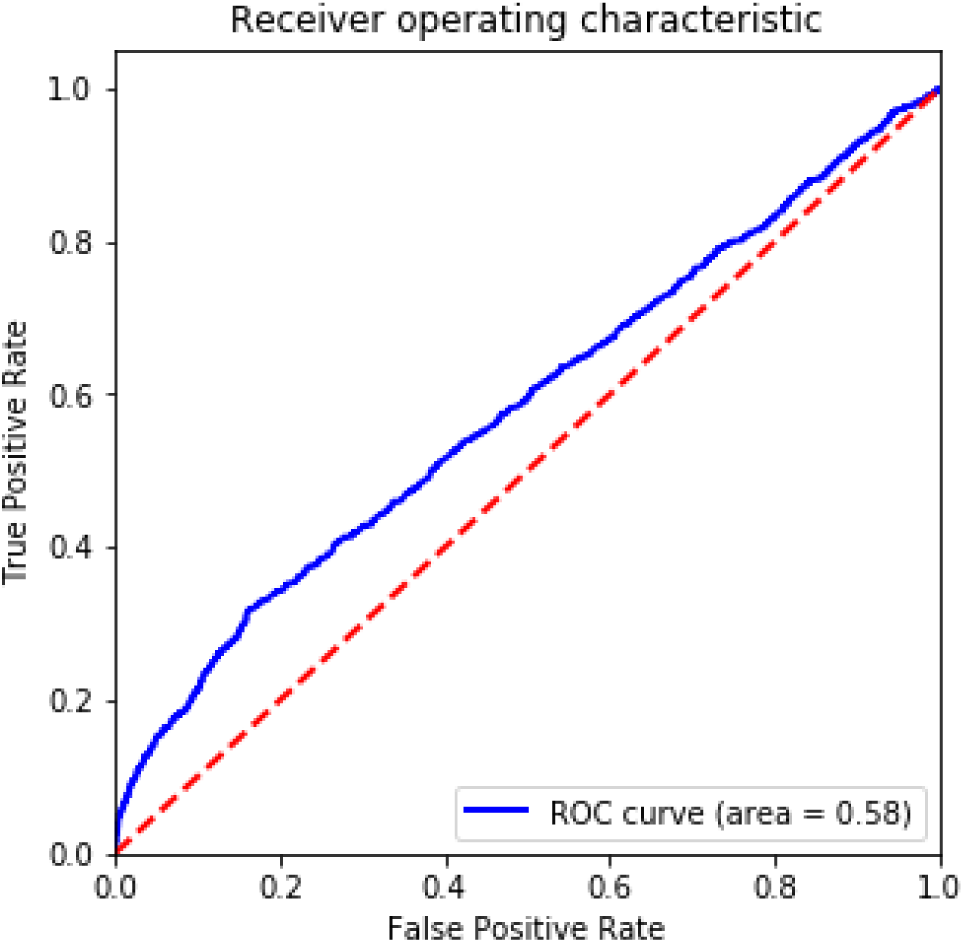
Receiver operating curve for the logistic regression model.

### 3.4 Feature Importance

Of the 52 most predictive features with non-zero weights learned from the logistic regression model, all features conveyed clinical meaningfulness in the application of early mortality prediction.

Age at initial diagnosis was the strongest predictor of early mortality, with a predictive feature weight of 0.157. Hispanic race was a significantly strong predictor of early mortality, with a weight of 0.074. The next strongest race-related predictor of early mortality is American Indian or Asian race (weight 0.0403).

Metastases were not strongly predictive of early mortality, with metastases to bone, brain, liver and lung resulting in weight of 0.0.

Certain histologies were highly predictive of early mortality, namely, complex mixed and stromal neoplasms (weight 0.107), epithelial neoplasms (weight 0.052), squamous cell neoplasms (weight 0.033), and fibroepithelial neoplasms (weight 0.022).

In terms of features strongly negatively predictive of early mortality, tumor size was the strongest negative predictor (weight −0.241), indicating that larger tumor sizes were negatively correlated with early mortality. Various histological subtypes were strongly negatively predictive of early mortality: specialized gonadal neoplasms (weight −0.081), basal cell neoplasms (weight −0.076), nevi and melanomas (weight −0.062), and germ cell neoplasms (weight −0.046).

Interestingly, white race was strongly negatively correlated with early mortality (weight −0.032).

Predictive features and their relative weights are visualized in terms of predicted weights in Figure 4.

**Figure 4:**
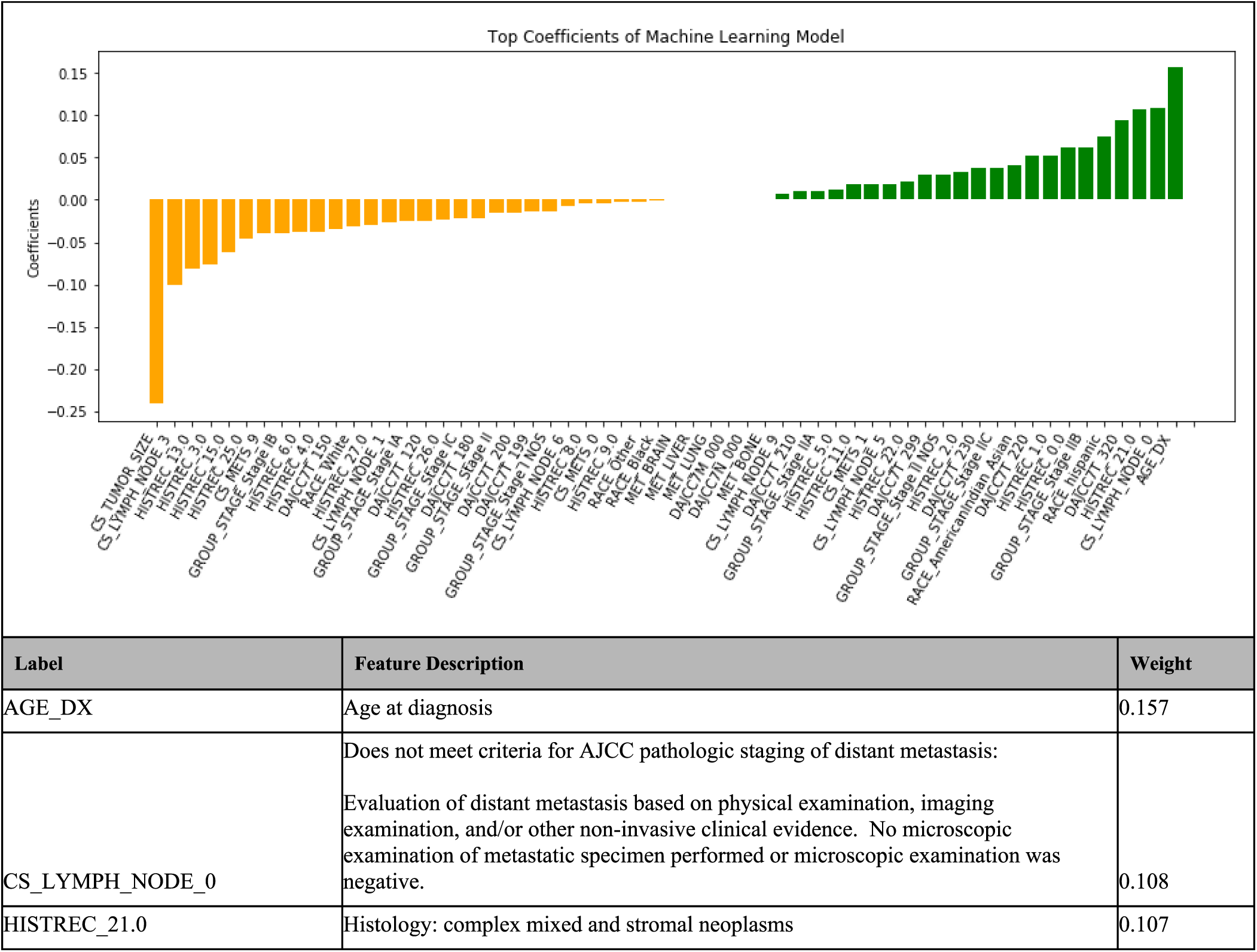
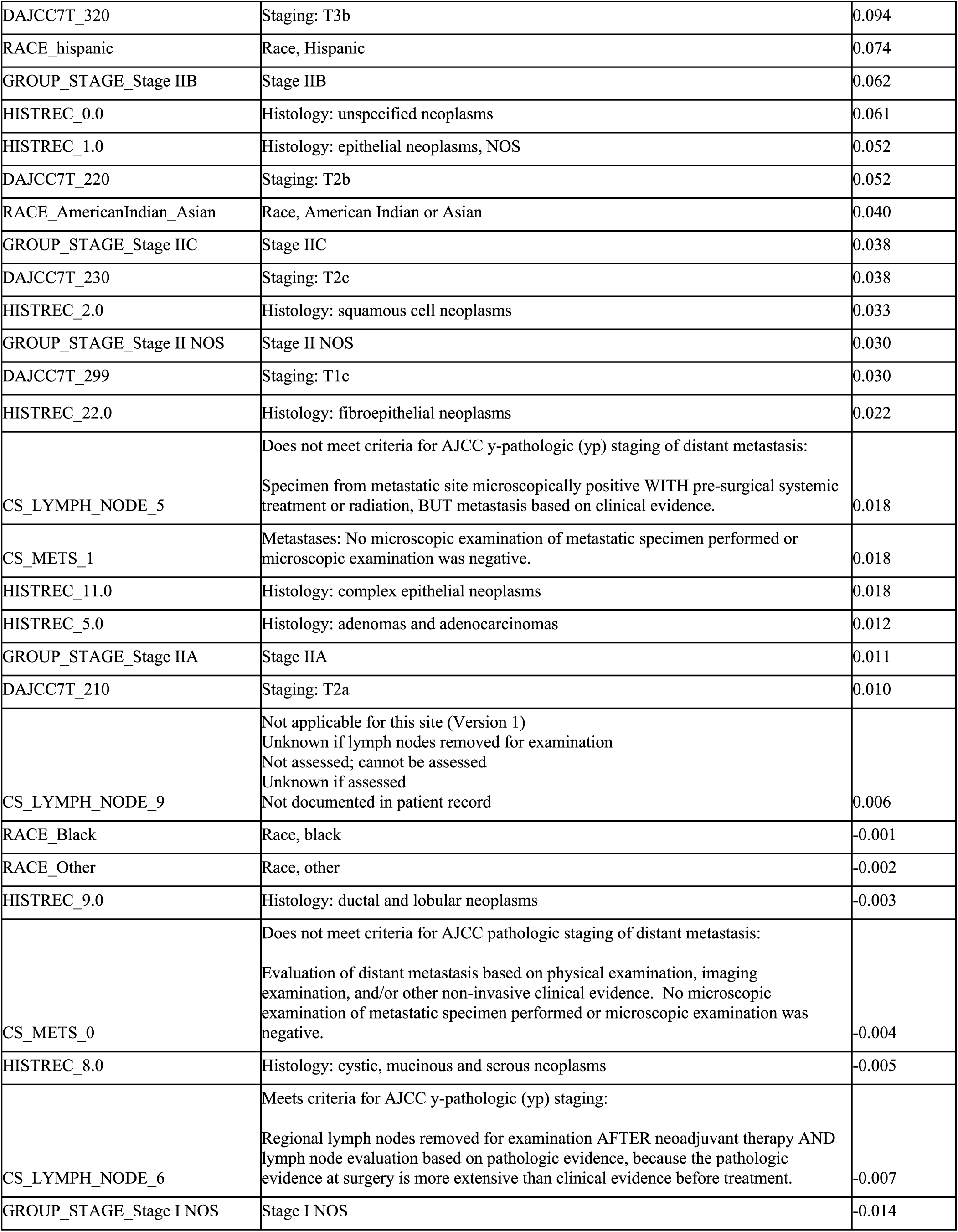
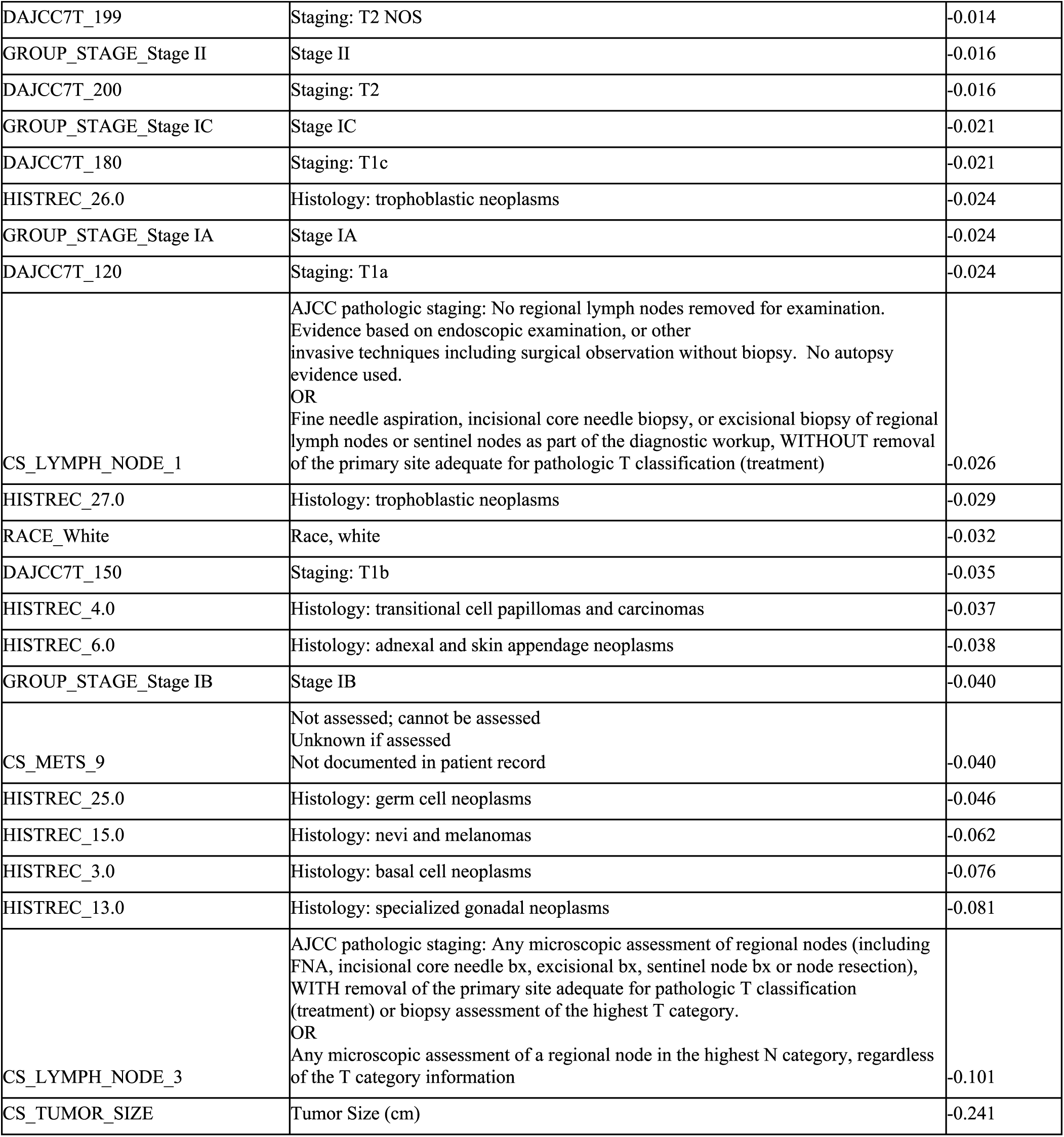
top most predictive features, including all features with a positive weight or negative weight learned from the model. Positive weights indicate positive correlation with early mortality, while negative weights indicate negative correlation with early mortality.

## 4. DISCUSSION

A machine learning model was developed to predict early mortality in patients with ovarian cancer, using an end point of less than 22 months, equivalent to the 25h percentile of overall survival in patients with stage I and stage II ovarian cancer.

The most predictive features were arrived at via extraction of weights learned from the logistic regression model. It is important to note that the most predictive features were determined from feature coefficients in the model. In our logistic regression model, feature importances were interpreted based on coefficients learned from the model. In clinical applications, interpretability is important for downstream usage of the model in personalized treatment plans for patients. Methods such as LIME(Ribeiro, Singh, and Guestrin 2016) have been successfully used in healthcare applications including prediction of mortality in ICU patients(Katuwal and Chen 2016).

It is important to note that there are methods for identifying risk factors as predictive of mortality, such as Cox proportional hazards model. We did not implement a Cox model in this scenario because the problem was setup as a prediction problem.

It is interesting to note several interesting trends with respect to the predictive features identified. Higher age was correlated with early mortality, as well as patients whose histology are of the subtypes of mplex mixed and stromal neoplasms (weight 0.107), epithelial neoplasms (weight 0.052), squamous cell neoplasms (weight 0.033), and fibroepithelial neoplasms (weight 0.022). On the other hand, patients’ whose primary histology were specialized gonadal neoplasms (weight −0.081), basal cell neoplasms (weight −0.076), nevi and melanomas (weight −0.062), and germ cell neoplasms (weight −0.046) were correlated with lower early mortality. Interestingly, tumor size, stage T3b, T2b, T2c stages were positively correlated with early mortality, while tumors of stages T1a and T1b were negatively correlated with early mortality. Patients with tumors of stage II, IA and IB were negatively correlated with early mortality. However, despite smaller tumor size being predictive of early mortality, metastases were not strongly predictive of early mortality.

We suggest 4 areas for future work. First, given the rare prevalence of ovarian cancer in the general population, we suggest thorough investigation of biomarkers associated with ovarian cancer. Biomarkers could include genomic, metabolomic, or microbiomic biomarkers. One example of a biomarker is circulating tumor DNA (ctDNA), which has been implicated in prognosis of neoplasms such as non Hodgkin’s lymphomas including follicular lymphoma(Delfau-Larue et al. 2018; Spina et al. 2018). Second, we suggest studies assessing the effectiveness of the model in a prospective setting. Third, we suggest exploration from public tumor registrics such as the Cancer Genome Atlas(Cancer Genome Atlas Research Network et al. 2013), the Catalogue of Somatic Mutations in Cancer(Forbes et al. 2011; Griffith et al. 2017), and the OncoKB Precision Oncology Knowledge Base (Ford 2017). Finally, we suggest unsupervised learning approaches to further characterize ovarian cancer. Approaches based on tensor factorization (Wang et al. 2015; Ho, Ghosh, and Sun 2014; Ho et al. 2014; Luo, Wang, and Szolovits 2017; Perros et al. 2017, 2018)may prove to be useful for uncovering distinct phenotypic subtypes of disease, as demonstrated in heterogeneous diseases such as heart failure.

## 5. CONCLUSION

We developed a machine learning model that predicts early mortality in patients with malignant neoplasm of the left testis with 76.1% accuracy with AUC 0.621, and is able to identify clinical features predictive of early mortality. Future work should include integration of additional data sources, as well as explore temporal modeling strategies to account for clinical changes over time.

## Data Availability

Public data were used.

https://seer.cancer.gov/

## Footnotes

1 See NCI SEER definition: https://training.seer.cancer.gov/schema/rp_ureter/reg_ln_eval.html

2 See NCI SEER definition: https://seer.cancer.gov/tools/ssm/2018-Summary-Stage-Manual.pdf

## REFERENCES

Cancer Genome Atlas Research Network, John N. Weinstein, Eric A. Collisson, Gordon B. Mills, Kenna R. Mills Shaw, Brad A. Ozenberger, Kyle Ellrott, Ilya Shmulevich, Chris Sander, and Joshua M. Stuart. 2013. “The Cancer Genome Atlas Pan-Cancer Analysis Project.” Nature Genetics 45 (10): 1113–20.

Cheng, Feixiong, and Zhongming Zhao. 2014. “Machine Learning-Based Prediction of Drug–drug Interactions by Integrating Drug Phenotypic, Therapeutic, Chemical, and Genomic Properties.” Journal of the American Medical Informatics Association: JAMIA 21 (e2): e278–86.

Chen, Robert, Vikas Kumar, Natalie Fitch, Jitesh Jagadish, Lifan Zhang, William Dunn, and Duen Horng Chau. 2015. “explICU: A Web-Based Visualization and Predictive Modeling Toolkit for Mortality in Intensive Care Patients.” Conference Proceedings:… Annual International Conference of the IEEE Engineering in Medicine and Biology Society. IEEE Engineering in Medicine and Biology Society. Conference 2015: 6830–33.

Chen, Robert, Walter F. Stewart, Jimeng Sun, Kenney Ng, and Xiaowei Yan. 2019. “Recurrent Neural Networks for Early Detection of Heart Failure From Longitudinal Electronic Health Record Data: Implications for Temporal Modeling With Respect to Time Before Diagnosis, Data Density, Data Quantity, and Data Type.” Circulation. Cardiovascular Quality and Outcomes 12 (10): e005114.

Chen, Robert, Hang Su, Mohammed Khalilia, Sizhe Lin, Yue Peng, Tod Davis, Daniel A. Hirsh, et al. 2015. “Cloud-Based Predictive Modeling System and Its Application to Asthma Readmission Prediction.” AMIA... Annual Symposium Proceedings / AMIA Symposium. AMIA Symposium 2015 (November): 406–15.

Choi, Edward, Andy Schuetz, Walter F. Stewart, and Jimeng Sun. 2017. “Using Recurrent Neural Network Models for Early Detection of Heart Failure Onset.” Journal of the American Medical Informatics Association: JAMIA 24 (2): 361–70.

Delfau-Larue, Marie-Hélène, Axel van der Gucht, Jehan Dupuis, Jean-Philippe Jais, Isabelle Nel, Asma Beldi-Ferchiou, Salma Hamdane, et al. 2018. “Total Metabolic Tumor Volume, Circulating Tumor Cells, Cell-Free DNA: Distinct Prognostic Value in Follicular Lymphoma.” Blood Advances 2 (7): 807–16.

Desautels, Thomas, Ritankar Das, Jacob Calvert, Monica Trivedi, Charlotte Summers, David J. Wales, and Ari Ercole. 2017. “Prediction of Early Unplanned Intensive Care Unit Readmission in a UK Tertiary Care Hospital: A Cross-Sectional Machine Learning Approach.” BMJ Open 7 (9): e017199.

Forbes, Simon A., Nidhi Bindal, Sally Bamford, Charlotte Cole, Chai Yin Kok, David Beare, Mingming Jia, et al. 2011. “COSMIC: Mining Complete Cancer Genomes in the Catalogue of Somatic Mutations in Cancer.” Nucleic Acids Research 39 (Database issue): D945–50.

Ford, James M. 2017. “Precision Oncology: A New Forum for an Emerging Field.” JCO Precision Oncology. https://doi.org/10.1200/po.16.00048.

Griffith, Malachi, Nicholas C. Spies, Kilannin Krysiak, Joshua F. McMichael, Adam C. Coffman, Arpad M. Danos, Benjamin J. Ainscough, et al. 2017. “CIViC Is a Community Knowledgebase for Expert Crowdsourcing the Clinical Interpretation of Variants in Cancer.” Nature Genetics 49 (2): 170–74.

Hankey, B. F., L. A. Ries, and B. K. Edwards. 1999. “The Surveillance, Epidemiology, and End Results Program: A National Resource.” Cancer Epidemiology, Biomarkers & Prevention: A Publication of the American Association for Cancer Research, Cosponsored by the American Society of Preventive Oncology 8 (12): 1117–21.

Hastie, Trevor, Robert Tibshirani, and Jerome Friedman. 2009. The Elements of Statistical Learning: Data Mining, Inference, and Prediction, Second Edition. Springer Science & Business Media.

Ho, Joyce C., Joydeep Ghosh, Steve R. Steinhubl, Walter F. Stewart, Joshua C. Denny, Bradley A. Malin, and Jimeng Sun. 2014. “Limestone: High-Throughput Candidate Phenotype Generation via Tensor Factorization.” Journal of Biomedical Informatics 52 (December): 199–211.

Ho, Joyce C., Joydeep Ghosh, and Jimeng Sun. 2014. “Marble: High-Throughput Phenotyping from Electronic Health Records via Sparse Nonnegative Tensor Factorization.” In Proceedings of the 20th ACM SIGKDD International Conference on Knowledge Discovery and Data Mining, 115–24. KDD ’14. New York, NY, USA: Association for Computing Machinery.

Johnson, Alistair E. W., Mohammad M. Ghassemi, Shamim Nemati, Katherine E. Niehaus, David Clifton, and Gari D. Clifford. 2016. “Machine Learning and Decision Support in Critical Care.” Proceedings of the IEEE. https://doi.org/10.1109/jproc.2015.2501978.

Kamat, Alisha, Ting Jin, So Yeon Min, Flaminia Talos, Jonas Almeida, and Daifeng Wang. 2018. “Interpretable Machine Learning Approach Reveals Developmental Gene Expression Biomarkers for Cancer Patient Outcomes at Early Stages.” Proceedings of the 2018 ACM International Conference on Bioinformatics, Computational Biology, and Health Informatics - BCB ’18. https://doi.org/10.1145/3233547.3233619.

Kaplan, E. L., and Paul Meier. 1958. “Nonparametric Estimation from Incomplete Observations.” Journal of the American Statistical Association 53 (282): 457–81.

Katuwal, Gajendra Jung, and Robert Chen. 2016. “Machine Learning Model Interpretability for Precision Medicine.” *arXiv [q-bio.QM]*. arXiv. http://arxiv.org/abs/1610.09045.

Luo, Yuan, Fei Wang, and Peter Szolovits. 2017. “Tensor Factorization toward Precision Medicine.” Briefings in Bioinformatics 18 (3): 511–14.

Makino, Masaki, Ryo Yoshimoto, Masaki Ono, Toshinari Itoko, Takayuki Katsuki, Akira Koseki, Michiharu Kudo, et al. 2019. “Artificial Intelligence Predicts the Progression of Diabetic Kidney Disease Using Big Data Machine Learning.” Scientific Reports 9 (1): 11862.

Ng, Kenney, Steven Steinbuhl, Christopher deFilippi, Sanjoy Dey, and Walter Stewart. 2017. “Early Detection of Heart Failure Using Electronic Health Records: Practical Implications for Time Before Diagnosis, Data Diversity, Data Quantity, and Data Density.” Journal of Patient-Centered Research and Reviews. https://doi.org/10.17294/2330-0698.1523.

Perros, Ioakeim, Evangelos E. Papalexakis, Haesun Park, Richard Vuduc, Xiaowei Yan, Christopher Defilippi, Walter F. Stewart, and Jimeng Sun. 2018. “SUSTain: Scalable Unsupervised Scoring for Tensors and Its Application to Phenotyping.” In Proceedings of the 24th ACM SIGKDD International Conference on Knowledge Discovery & Data Mining, 2080–89. KDD ’18. New York, NY, USA: Association for Computing Machinery.

Perros, Ioakeim, Evangelos E. Papalexakis, Fei Wang, Richard Vuduc, Elizabeth Searles, Michael Thompson, and Jimeng Sun. 2017. “SPARTan: Scalable PARAFAC2 for Large & Sparse Data.” In Proceedings of the 23rd ACM SIGKDD International Conference on Knowledge Discovery and Data Mining, 375–84. KDD ’17. New York, NY, USA: Association for Computing Machinery.

Rajkomar, Alvin, Eyal Oren, Kai Chen, Andrew M. Dai, Nissan Hajaj, Michaela Hardt, Peter J. Liu, et al. 2018. “Scalable and Accurate Deep Learning with Electronic Health Records.” NPJ Digital Medicine 1 (May): 18.

Rasmy, Laila, Yonghui Wu, Ningtao Wang, Xin Geng, W. Jim Zheng, Fei Wang, Hulin Wu, Hua Xu, and Degui Zhi. 2018. “A Study of Generalizability of Recurrent Neural Network-Based Predictive Models for Heart Failure Onset Risk Using a Large and Heterogeneous EHR Data Set.” Journal of Biomedical Informatics 84 (August): 11–16.

Ribeiro, M. T., S. Singh, and C. Guestrin. 2016. “‘ Why Should I Trust You?’ Explaining the Predictions of Any Classifier.” Proceedings of the 22nd ACM. https://dl.acm.org/doi/abs/10.1145/2939672.2939778.

Spina, Valeria, Alessio Bruscaggin, Annarosa Cuccaro, Maurizio Martini, Martina Di Trani, Gabriela Forestieri, Martina Manzoni, et al. 2018. “Circulating Tumor DNA Reveals Genetics, Clonal Evolution, and Residual Disease in Classical Hodgkin Lymphoma.” Blood 131 (22): 2413–25.

Wang, Yichen, Robert Chen, Joydeep Ghosh, Joshua C. Denny, Abel Kho, You Chen, Bradley A. Malin, and Jimeng Sun. 2015. “Rubik: Knowledge Guided Tensor Factorization and Completion for Health Data Analytics.” KDD: Proceedings / International Conference on Knowledge Discovery & Data Mining. International Conference on Knowledge Discovery & Data Mining 2015 (August): 1265–74.

Yu, Chao, Jiming Liu, and Shamim Nemati. 2019. “Reinforcement Learning in Healthcare: A Survey.” *arXiv [cs.LG]*. arXiv. http://arxiv.org/abs/1908.08796.

